# Implementation of *CYP2D6* copy-number imputation panel and frequency of key pharmacogenetic variants in Finnish individuals with a psychotic disorder

**DOI:** 10.1101/2020.11.13.20227058

**Authors:** K. Häkkinen, JI. Kiiski, M. Lähteenvuo, T. Jukuri, K. Suokas, J. Niemi-Pynttäri, T. Kieseppä, T. Männynsalo, A. Wegelius, W. Haaki, K. Lahdensuo, R. Kajanne, MA. Kaunisto, A. Tuulio-Henriksson, O. Kampman, J. Hietala, J. Veijola, J. Lönnqvist, E. Isometsä, T. Paunio, J. Suvisaari, E. Kalso, M. Niemi, J. Tiihonen, M. Daly, A. Palotie, AV. Ahola-Olli

## Abstract

**Purpose:** We constructed a *CYP2D6* copy-number imputation panel by combining copy-number information to GWAS chip data. In addition, we report frequencies of key pharmacogenetic variants in individuals with a psychotic disorder from the genetically bottle-necked population of Finland.

**Methods:** We combined GWAS chip and *CYP2D6* copy-number variation (CNV) data from the Breast Cancer Pain Genetics study (BrePainGen) to construct an imputation panel (N=902) for *CYP2D6* CNV. The resulting data set was used as a *CYP2D6* CNV imputation panel in 9,262 non-related individuals passing genotype data quality control procedures. The panel performance was evaluated by genotyping the CNV from a subset (N=297) of SUPER-Finland participants.

**Results:** *CYP2D6* CNV was imputed correctly in 272 (92%) individuals. Sensitivity and specificity for detecting a duplication were 0.986 and 0.946, respectively. Sensitivity and specificity for detecting a deletion using imputation were 0.886 and 0.966, respectively. Based on imputation, the frequency of a *CYP2D6* duplication and deletion in the whole SUPER-Finland sample with 9,262 non-related individuals passing quality control were 8.5% and 2.7%, respectively. We confirm the higher frequency of CYP2D6 ultrarapid metabolizers in Finland compared with non-Finnish Europeans. Additionally, we confirm a 21-fold enrichment of the *UGT1A1* decreased function variant rs4148323 (also known as 211G>A, G71R or *UGT1A1*\*6) in Finland compared with non-Finnish Europeans. Similarly, the *NUDT15* variant rs116855232 was highly enriched in Finland.

**Conclusion:** Our results demonstrate that imputation of *CYP2D6* CNV is possible. The methodology is not accurate enough to be used in clinical decision making, but it enables studying *CYP2D6* in large biobanks with genome-wide data. In addition, it allows for researchers to recontact patients with certain pharmacogenetic variations through biobanks. We show that bottle-necked populations may have pharmacogenetically important variants with allele frequencies very different from the main ancestral group. Future studies should assess whether these differences are large enough to cause clinically significant changes in trial results across different ancestral groups.

## Introduction

Pharmacogenetics is a research field studying how interindividual genetic differences contribute to drug efficacy and safety, aiding physicians in drug selection and dose adjustment^1,2^. Variants in *CYP2D6* and *CYP2C19* genes have been shown to affect the metabolism of antidepressants, antipsychotics, analgesics such as codeine and tramadol, the antiplatelet agent clopidogrel and antiarrhythmic drugs, for example^3,4,5,6,7,8^. Although involved in the metabolism of 25% of all available drugs, research of the *CYP2D6* locus at 22q13.2 has been limited by its complexity^9,10^. In addition to single nucleotide polymorphisms (SNPs), *CYP2D6* copy-number variations (CNVs), such as duplications and deletions, also contribute to the metabolic activity of the CYP2D6 enzyme. The effects of SNPs and CNVs on the metabolic activity of CYP2D6 can be summarized by calculating an activity score^11^. The frequency of the *CYP2D6* gene duplication is highly dependent on the population^12,13,14,15,16^. The effects of genetic variants on drug metabolism are large enough to cause the Food and Drug Administration (FDA) and the European Medicines Agency (EMA) to add pharmacogenetic information on drug labels^17,18,19^ and the American Psychiatric Association to inform clinicians on pharmacogenetics in recently updated schizophrenia guidelines^20^. Whether the difference in allele frequency explains why different ethnicities respond to certain drugs differently is difficult to estimate, as there are other cultural and socio-economic factors at play as well^21^.

Genotype imputation has been successfully used in genome-wide association studies (GWAS) to acquire data on ungenotyped markers, facilitate fine-mapping, and boost power in association studies^22^. The method relies on the haplotype structure of the human genome. Haplotypes are block-like regions of DNA. Within these blocks certain variants tend to co-occur allowing for separation of maternally and paternally inherited variants. Information of which variants are located on the same chromosome is needed to accurately impute missing variants. This information also has an important role in pharmacogenetics since it allows to infer whether, for example, two loss-of-function variants in the same gene are located on the same chromosome or not. If they resided on different homologous chromosomes, the individual would be a complete knock-out for the pertinent gene. On the other hand, if these two variants resided on the same chromosome, the individual would still carry one functional copy of this gene resulting in a pharmacogenetically different situation. The process of computationally separating the variants to different chromosomes is called phasing^23^. Usually, only SNPs or short indels are imputed, but since *CYP2D6* duplications and deletions tend to co-occur with certain SNPs, we hypothesized that existing imputation algorithms could be exploited to impute *CYP2D6* CNVs to a large set of GWAS chip-genotyped individuals. Usually, CNVs are genotyped with real-time PCR as *CYP2D6* CNVs are too small (4 kilobases) to be detected from GWAS chip signal intensity data. Successful imputation would allow for genotyping only a subset of the sample (a few hundred individuals), after which the CNV carrier status could be predicted *in silico* for hundreds of thousands of individuals. This would result in cost-effective pharmacogenetic analyses in biobanks and other large GWAS chip-genotyped samples, such as FinnGen (www.finngen.fi).

Next generation sequencing has become increasingly popular during the recent years. Several algorithms already exist to predict *CYP2D6* carrier status from sequencing depth, which is a measure of overlapping reads in a given region of the genome. Duplicated genome regions tend to have a higher sequencing depth compared with the regions of the reference genotype as more reads are generated from the duplicated region. However, sequencing data are available from smaller sample sets compared with GWAS chip data. Furthermore, *CYP2D6* CNV algorithms relying on sequencing depth tend to provide discordant CNV calls^24^.

Due to the small founder population, bottleneck effects and genetic drift, the Finnish population is enriched for certain low frequency (0.5-5%) variants, including coding and loss-of-function variants^25,26^. Thus, our secondary aim in this work was to assess whether the unique population history has caused alterations in the frequency of pharmacogenetic variants compared with other European populations.

## Material and methods

A flow chart of the study protocol is provided in Figure 1.

**Figure 1.**
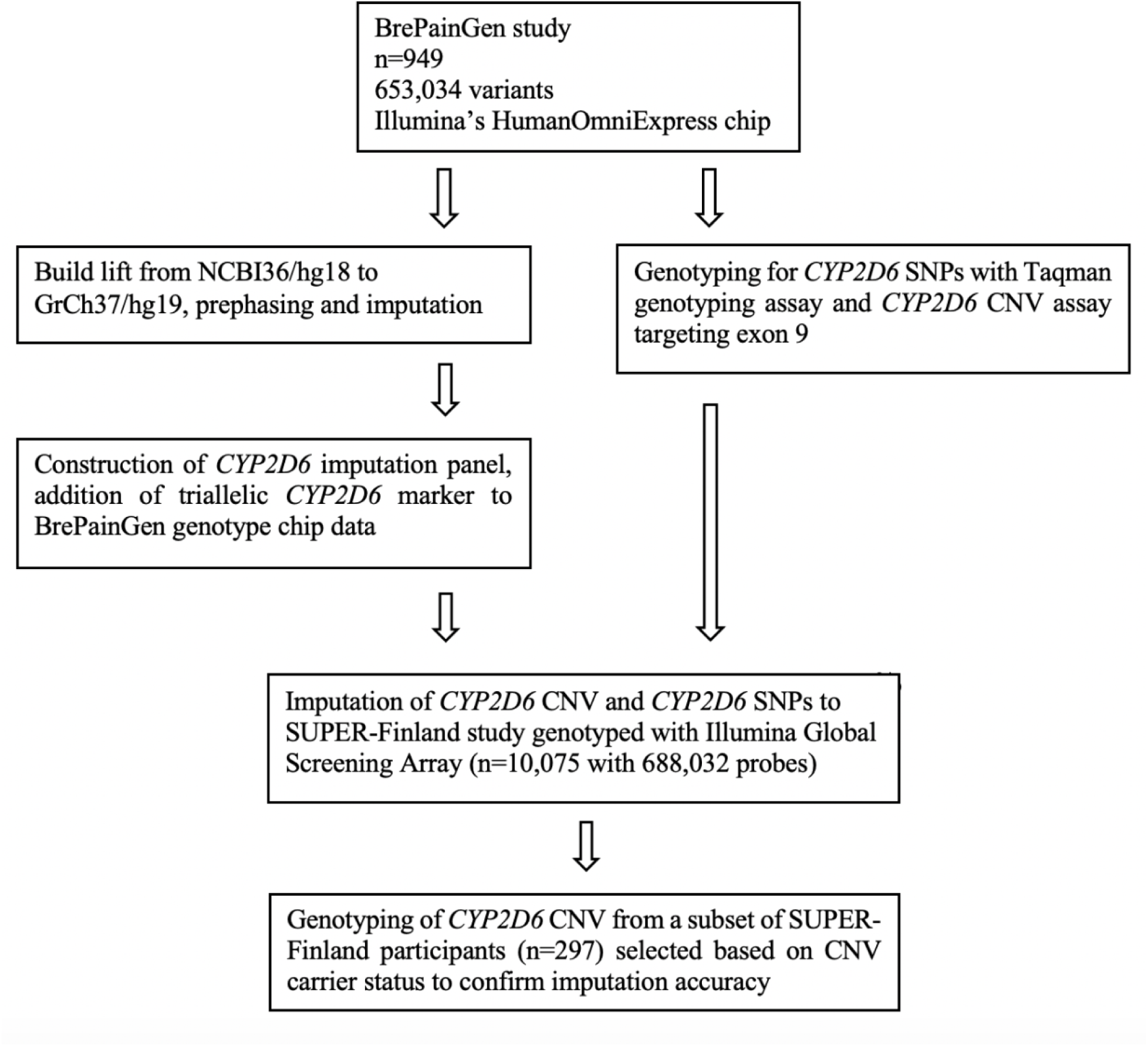
A flow chart of study protocol.

### SUPER-Finland study

The SUPER-Finland study recruited 10,474 participants aged >18 with a severe mental disorder between the years 2016-2018 from Finland. Subjects were recruited from in- and outpatient psychiatric, general care, and housing units with a diagnosis of a schizophrenia spectrum psychotic disorder (ICD-10 codes F20, F22-29), bipolar disorder (F30, F31) or major depressive disorder with psychotic features (F32.3 and F33.3). As Finland contains internal genetic subisolates^27,28^, special care was taken to ensure wide coverage of known isolate areas.

Blood samples were drawn from participants for DNA extraction (2x Vacutainer EDTA K2 5/4 ml, BD). When venipuncture was not possible, saliva sample (DNA OG-500, Oragene) was collected for DNA extraction. All samples were frozen (−20^°^C) on site within 60 min of sampling and sent to the THL (Finnish Institute for Health and Welfare) Biobank within 3 months for long term storage in −185°C. DNA was extracted from EDTA-blood tubes using PerkinElmer Janus chemagic 360i Pro Workstation with the CMG-1074 kit. After incubation in +50 °C, o/n DNA was extracted from saliva samples with Chemagen Chemagic MSM I robot with CMG-1035-1 kit. DNA samples were shipped on dry ice to the Broad Institute of MIT and Harvard, Boston Cambridge, Massachusetts, USA for genotyping and sequencing.

ICD-code diagnosis and disease duration (as years from receiving the diagnosis until recruitment) of the SUPER-Finland participants were extracted from The Care Register for Health Care^29^.

### Genotyping and sequencing

10,075 SUPER-Finland individuals were genotyped with Illumina Global Screening Array containing 688,032 probes. Genotyping was performed at Broad Institute in Cambridge, Massachusetts, USA. Subjects with a genotyping success rate < 90% and discordance between reported gender and genotyped sex were excluded. After this, variants with over 90% of missing genotype calls and related samples using pi-hat cut-off of 0.15 were excluded. Variants deviating from Hardy-Weinberg equilibrium were excluded (P < 1×10^−8^). Samples with low or excess heterozygocity (±3*SD* from sample mean) were excluded. Imputation was performed using the FinnGen-style imputation protocol as described in protocols.io (https://www.protocols.io/view/genotype-imputation-workflow-v3-0-nmndc5e?version_warning=no). Sequencing Initiative Suomi (SISu) v2 panel was used as the imputation reference^30^.

### Construction and validation of CYP2D6 CNV imputation panel

A *CYP2D6* CNV imputation panel was constructed using data from the Breast Cancer Pain Genetics Study (BrePainGen^31,32^). The Finnish BrePainGen consisted of 1000 patients recruited between 2006-2010, who underwent surgery for breast cancer at the Helsinki University Hospital. The BrePainGen subjects were genotyped with HumanOmniExpress-12v1_H chip manufactured by Illumina. Before quality control, we had data consisting of 949 samples and 733,202 probes. Probes with more than 3 % missing data were excluded (222 probes failed). After this, samples with over 3% missingness rate (0 excluded) were filtered. Variants with minor allele frequency below 0.5% were excluded (63918 variants excluded). Subsequently, variants with Hardy-Weinberg p-value < 1×10E-6 were excluded (10,857 fails). Sex check was not performed as all samples were from female study participants. In the heterozygosity check, 14 samples failed. Five samples were excluded due to relatedness. Based on MDS plots, 4 individuals were excluded. The final data set consisted of 653,034 variants and 926 samples. The genotyping rate was 0.9968. The initial dataset was lifted from NCBI36/hg18 to GrCh37/hg19 using Will Rayner’s method (https://www.well.ox.ac.uk/~wrayner/strand/). *CYP2D6* CNV was genotyped separately with real-time PCR^32^. Data on CNV were converted with R to Plink ped- and map-file format. This was then joined to the quality controlled GWAS chip data with Plink’s (version 1.9) --merge option. Next, the data were pre-phased with Eagle^33,34^ version 2.4 and imputed with Beagle^35^ version 4.1 software. The software code for CNV imputation pipeline is available upon reasonable request.

### Genotyping of CYP2D6 CNV in a subset of SUPER-Finland participants

To confirm the copy-number imputation results, 297 SUPER-Finland subjects were selected for *CYP2D6* CNV genotyping based on CNV and SNP imputation results to guarantee a sufficient number of deletion and duplication carriers in addition to *41, and *10 carriers in the validation set. Samples with possible fusion gene were excluded (n=9). Real-time PCR genotyping was performed on QuantStudio™ 12K Flex system (Thermo Fisher Scientific, Waltham, MA) with three TaqMan^®^ Copy Number Assays: Hs00010001_cn targeting exon 9, hs04502391_cn targeting intron 6, and hs04083572_cn targeting intron 2 of *CYP2D6* gene. Four replicates of each sample were genotyped. The reaction volume was 10 μl and RNase P was used as a reference assay. The copy-number for each sample was calculated with the CopyCaller^™^ Software (Applied Biosystems®) according to the manufacturer’s instructions.

### Haplotype construction according to CPIC guidelines

To predict pharmacogenetic phenotypes from genotype, functional annotations described in The Clinical Pharmacogenetics Implementation Consortium (CPIC) guidelines were followed for variants in the *CYP2C9*^36^, *CYP2C19*^37^, *CYP2D6*^38^, *DPYD*^39^, *NUDT15*^40^, *SLCO1B1*^41^, *TPMT*^40^ and *UGT1A1*^42^ genes. Variants included in the phenotype prediction from the SUPER-Finland data are described in Supplement Table 1. If an individual did not carry any of these variants, but had been genotyped, the predicted phenotype was defaulted to normal.

### Statistical analyses

To provide estimates for the accuracy of the imputation method, we calculated sensitivity, specificity, negative predictive value (NPV) and positive predictive value (PPV), as described earlier^43^.

To calculate the differences in the geographical distribution of the predicted pharmacogenetic phenotypes, we used Fisher’s test. All statistical analyses were performed in R version 3.5.1^44^.

### Ethics statement

The SUPER-Finland study was given a favorable ethics statement (202/13/03/00/15) by the Coordinating Ethics Committee of the The Hospital District of Helsinki and Uusimaa (HUS). The BrePainGen study was approved by the Coordinating Ethics Committee (136/E0/2006) and the Ethics Committee of the Department of Surgery (Dnro 148/E6/05) of the Hospital District of Helsinki and Uusimaa (HUS). Written informed consent was obtained from each participant prior to inclusion in both of the studies.

## Results

9,262 non-related individuals participating in the SUPER-Finland study passed genotype quality control steps and thus had imputed genotype data available, 49.6% of these samples were female. The descriptive statistics of the sample are shown in Table 1.

**Table 1.** Characteristics of study participants (total n= 9262) in SUPER-Finland.

|  |  |
| --- | --- |
| **Gender % (n)** |  |
| Women | 49.6 (4589) |
| Men | 50.4 (4660) |
| Unknown | 0.1(13) |
| **Age mean (SD)** | 46.6 (14.8) |
| **Recruitment region % (n)** |  |
| Helsinki district (Southern Finland) | 25.2 (2329) |
| Tampere district (Central Finland) | 24.8 (2298) |
| Kuopio district (Eastern Finland) | 20.4 (1886) |
| Oulu district (Northern Finland) | 18.7 (1736) |
| Turku district (Western Finland) | 10.9 (1013) |
| **Diagnosis % (n)** |  |
| Schizophrenia | 55.4 (5117) |
| Bipolar disorder (I & II) | 15.4 (1419) |
| Other psychosis | 10.1 (934) |
| Schizoaffective disorder | 9.1 (837) |
| Psychotic depression | 5.2 (475) |
| Other mental disorder | 4.8 (447) |
| Unknown | 0.4 (33) |
| **Disease duration (years) mean (SD)** |  |
| Schizophrenia | 21.9 (12.8) |
| Bipolar disorder (I & II) | 13.7 (9.0) |
| Other psychosis | 15.2 (11.6) |
| Schizoaffective disorder | 21.8 (12.1) |
| Psychotic depression | 14.2 (9.0) |

### CYP2D6 CNV imputation panel

The performance of the *CYP2D6* CNV imputation panel was evaluated by genotyping CNVs with real-time PCR from 297 SUPER-Finland subjects. Sensitivity, specificity, PPV and NPV are reported in Supplemental Table 2. The contingency table of imputed and PCR-genotyped *CYP2D6* CNVs in SUPER-Finland is presented in Table 2. Because subjects were selected based on expected copy-number for validation, NPV and PPV do not represent the situation in the general population or in the patients with psychosis as PPV and NPV are dependent on CNV frequency. The imputation method was able to identify all except one true duplication carrier as having CYP2D6 duplication but it misclassified additional 12 subjects as having CN=3 although these 12 subjects did not carry duplications. This results in high sensitivity (0.986) for detecting duplications. *CYP2D6* deletions were imputed correctly for 31 individuals. The method misidentified 4 individuals carrying a deletion as either having a normal copy-number or carrying a duplication (false negatives). Overall, the method showed good accuracy as *CYP2D6* CNV was imputed correctly for 272 (92%) individuals.

**Table 2.** A contingency table of imputed and real-time PCR genotyped *CYP2D6* copy-number (CN) in SUPER-Finland.

|  |  | Imputed copy-number |  |  |
| --- | --- | --- | --- | --- |
|  |  | CN=1 | CN=2 | CN=3 |
| Genotyped<br>copy-number | CN=1 | 31 | 3 | 1 |
|  | CN=2 | 9 | 168 | 11 |
|  | CN=3 | 0 | 1 | 73 |

### Frequencies of key pharmacogenetic variants in SUPER-Finland

Predicted phenotypes and prevalence of *CYP2C9, CYP2C19, CYP2D6, DPYD, NUDT15, SLCO1B1, TPMT, UGT1A1* phenotypes in SUPER-Finland (total n=9,262) are described in Table 3. Observed minor allele frequencies of the variants used in the phenotype prediction are included in Supplemental Table 1. Based on *CYP2D6* imputation, *CYP2D6* gene duplication occurred in 8.5% (n=791) and deletion in 2.7% (n=247) of the SUPER-Finland participants. A total of 26 individuals carried the duplication in both homologous chromosomes and two individuals had both gene copies deleted. When these structural re-arrangements were combined with SNPs and translated to predicted CYP2D6 phenotypes, we observed that 6.6% (n=607) of the participants were ultrarapid metabolizers (UMs), 62.7% (n=5811) were normal metabolizers (NMs), 27.5% (n=2545) were intermediate metabolizers (IMs) and 3.2% (n=299) were poor metabolizers (PMs).

**Table 3.** Predicted phenotype and prevalence of *CYP2C9, CYP2C19, CYP2D6, DPYD, NUDT15, SLCO1B1, TPMT* and *UGT1A1* in SUPER-Finland (total n=9,262).

| **GENE** | **PREDICTED PHENOTYPE** | **PREVALENCE % (N)** |
| --- | --- | --- |
| ***CYP2C9*** | Normal | 67.3 (6230) |
|  | Intermediate (AS 1.5) | 19.7 (1823) |
|  | Intermediate (AS 1.0) | 11.0 (1022) |
|  | Poor | 2.0 (187) |
| ***CYP2C19*** | Ultrarapid | 3.8 (355) |
|  | Rapid | 24.3 (2254) |
|  | Normal | 39.7 (3676) |
|  | Intermediate | 28.7 (2656) |
|  | Poor | 3.5 (321) |
| ***CYP2D6*** | Ultrarapid | 6.6 (607) |
|  | Normal | 62.7 (5811) |
|  | Intermediate | 27.5 (2545) |
|  | Poor | 3.2 (299) |
| ***DPYD*** | Normal | 92.9 (8604) |
|  | Intermediate (AS 1.5) | 2.6 (242) |
|  | Intermediate (AS 1) | 4.4 (403) |
|  | Poor (AS 0.5) | 0.05 (5) |
|  | Poor (AS 0) | 0.09 (8) |
| ***NUDT15*** | Normal | 96.3 (8915) |
|  | Intermediate | 3.6 (336) |
|  | Poor | 0.1 (11) |
| ***SLCO1B1*** | Normal | 62.6 (5795) |
|  | Decreased | 33.6 (3110) |
|  | Poor | 3.9 (357) |
| ***TPMT*** | Normal | 94.1 (8712) |
|  | Intermediate | 5.8 (539) |
|  | Poor | 0.1 (11) |
| ***UGT1A1*** | Normal | 31.8 (2949) |
|  | Intermediate | 48.8 (4522) |
|  | Poor | 19.3 (1791) |

AS; Activity Score.

The prevalence of CYP2C19 UMs was 3.8% (n=355). 24.3% (n=2254) were classified as RMs and 3.5% (n=321) as PMs, 39.7% (n=3676) were NMs and the remaining 28.7% (n=2656) were IMs. The predicted phenotypes for CYP2C9 were as follows: NM 67.3% (n=6230), IMs with 1.5 activity score 19.7% (n=1823), IMs with 1 activity score 11.0% (n=1022) and PMs 2.0% (n=187).

As the population history of Finland has created genetic subisolates within the country, we compared whether the prevalence of the pharmacogenetic phenotypes of CYP2D6 and CYP2C19 differ by recruitment center (Supplemental Tables 3 and 4). For CYP2D6 UMs, the largest absolute difference was observed between Kuopio and Oulu (5.0% vs 7.7%; OR 0.63; 95% CI 0.47-0.83; P < 8×10^−4^).

A *NUDT15* variant classified as having no function (rs116855232-T) was enriched in Finland for 6.5-fold (P = 2.09× 10^−181^) when comparing Finns and non-Finnish Europeans from GnomAD v2.1.1 (https://gnomad.broadinstitute.org)^45^. The minor allele frequency was 0.02 in GnomAD Finns as well as in SUPER-Finland (Supplemental Table 1) whereas it is 0.004 for Europeans in general.

A variant in *UGT1A1* gene encoding *6 haplotype (rs4148323-A) was enriched in Finland 22-fold (Supplemental Table 1). The minor allele frequency in Finns was 5% compared to 0.2% in non-Finnish Europeans.

## Discussion

We have shown that the *CYP2D6* copy-number can be reliably imputed for research purposes. The presented imputation method allows for imputation of the *CYP2D6* copy-number in large samples such as the UK Biobank and FinnGen. This in turn allows detailed cost-effective pharmacogenetic analyses, as these data sets have longitudinal drug prescription history available. Studying pharmacogenetics in psychiatric patients is important as many drugs used in psychiatry are metabolized through polymorphic CYP-enzymes and some of these drugs, such as antidepressants, exert their effects slowly compared with diabetes medication, for example^46,47^. Thus, a 4-8 weeks follow-up is recommended before switching for example an antidepressant to another^48^. Therefore, a lack of efficacy from the initial treatment may only be discovered weeks or months after the initiation of the treatment and may lead to a major slow-down in the patient’s recovery process. Evidence is already emerging that polymorphisms in genes encoding CYP enzymes are associated with clinically significant outcomes^49^. Interestingly, the drug concentrations and adverse drug reactions seem to be under strong monogenic or oligogenic control with only one or few loci associated with them (at least with current sample sizes) whereas many psychiatric disorders are extremely polygenic^50,51,52,53^. As accurate inference of metabolic activity of CYP2D6 cannot currently be done from GWAS chip data alone, the copy-number imputation method creates new opportunities for development towards and research of individualized medicine. When more data emerge, the pharmacogenetic decision algorithms can be then fine-tuned to reflect newest findings^54^. Hence, it is important that pharmacogenetic reports can be updated to maintain best possible dosing guidelines available for the patients.

For research use, the correlation between imputed and true copy-number is not required to be perfect if we are studying large cohorts, as even modest correlations will add information. To achieve similar statistical power for detecting an association using an imputed marker as opposed to using a directly genotyped one, the sample size must be increased by 5-13% for each 1% increase in imputation error^55^. Given that many pharmacogenetic studies conducted so far have a sample size below 2,000 individuals, statistical power gained through increasing sample size to biobank scale (500k samples) would overcome the power loss due to inaccuracy in imputation. The imputation inaccuracy can be passed to statistical models for examples as allelic dosages, which range between 0 and 2. The closer the dosage is to a round number (0, 1, or 2), the more accurate the imputation result is. Other measures, such as posterior probability, have also been used^22^. As some pharmacogenetic variants are rare, imputation might be even more accurate than direct genotyping of the variant of interest with a GWAS chip, as calling algorithms perform poorly on rare variants and thus they are usually excluded before imputation^56,57^.

Genetic variation in *CYP2D6* has been described and compared between Finns and other European populations by earlier smaller studies. The Finns have been shown to have a high frequency of *CYP2D6* duplications and UM phenotypes compared with the ancestral European population^15,58^. Here, we show a high frequency of CYP2D6 UMs among subjects with a psychotic disorder throughout the country. *CYP2D6* genotype has an effect on metabolism of antipsychotics, such as risperidone and aripiprazole. This effect is further reflected on the therapeutic failure rate during risperidone therapy, suggesting that genotyping could be used to guide dosing decisions^4^.

We confirm a high frequency of the *UGT1A1* variant rs4148323, also known as *UGT1A1*6*, in Finland. This variant has been linked to irinotecan toxicity in Biobank Japan^59^. According to GnomAD v2.1.1, the frequency of rs4148323-A in Europeans is only 0.2% but here we report, as seen earlier in a small sample size study^60^, that the frequency in Finns is 4.2% which means an approximately 21-fold enrichment. The study concerning irinotecan-treated patients from Biobank Japan demonstrated that 51 out of the 330 subjects with normal UGT1A1 metabolism experienced adverse drug reactions (15%) whereas 8 out of 15 subjects (53%) with rs4148323-AA experienced an adverse drug reaction. As rs4148323-A is rare among European subjects, the drug trials involving irinotecan might not have captured the increased risk related to this genotype in Finns. Another *UGT1A1* variant, linked to decreased irinotecan metabolism is *UGT1A1*28* or rs8175347^61^. The pertinent variant is also linked to Gilbert syndrome characterized by periods of intermittent icterus^62^. The variant inserts a seventh TA repeat sequence to already six TA repeats within *UGT1A1* thus decreasing the enzyme activity by 70%^63^. Since rs8175347 was not present in the SISu imputation panel, we used a proxy variant (rs887829) to estimate the frequency of *UGT1A1*28* in Finns. The proxy variant is in strong linkage with rs8175347 in Finns according to LDlink^64^ (r-squared 1.0) and also associated with Gilbert syndrome in FinnGen (https://www.finngen.fi/en/access_results). The proxy also shows an increased frequency among Finns compared with the rest of Europe, which leads to a high frequency of UGT1A1 poor metabolizers in Finland. This should be considered when planning to initiate irinotecan treatment for patients with Finnish ancestry. Finnish Biobank samples should be used to evaluate risks associated with irinotecan treatment as studies conducted elsewhere in Europe may not be generalizable to Finland.

*CYP2C19* genotypes have been shown to contribute to the metabolism of several antidepressants, clopidogrel, and proton pump inhibitors^3,37,65,66,67^. Subjects with poor or intermediate CYP2C19-mediated metabolism are at increased risk for recurrent myocardial infarction after percutaneous coronary angioplasty when treated with clopidogrel as the activation of the drug is insufficient^68,69^. CYP2C19 genotype is also associated with a failure of escitalopram treatment. Jukic et al. reported that 30.7% of CYP2C19 PMs discontinued escitalopram compared to discontinuation rates of 11.8%, 17.8%, and 28.9% in normal, rapid, and ultrarapid metabolizers, respectively^3^. Thus, subjects with an increased discontinuation rate make up 31.6 % of SUPER-Finland subjects, which is a clinically significant proportion of the patient base.

Recently, a new compound called siponimod, was introduced to markets in Europe and the USA for the treatment of multiple sclerosis^70^. The therapeutic dose of siponimod is dependent on the *CYP2C9* genotype. In addition to siponimod, the *CYP2C9* gene has a well-established role in determining the warfarin dose^71^. According to the manufacturer of siponimod, the drug is contraindicated in patients with the *CYP2C9* *3/*3 genotype (rs1057910-CC). In addition, dose adjustment is needed for *2/*3 and *1/*3 genotypes. Thus, genotyping is necessary before initiating the drug. According to our results, the drug is contraindicated in about 0.5% of Finns and 11.2% require dose adjustments.

*DPYD* variants have a large impact on the safety of the chemotherapeutic agents capecitabine and 5-fluorouracil. Based on the phenotype prevalence observed here, 7.1% (n=658) of individuals would require dose adjustment for these drugs according to the CPIC guidelines^39^.

Variations in *NUDT15* were recently introduced in the CPIC guidelines to help estimate azathioprine dose for the treatment of Crohn’s disease, for example^40^. When determining proper azathioprine dose, *NUDT15* variants are interpreted together with *TPMT* variants. According to CPIC’s decision algorithm, the dose is determined by the gene the function of which is most severely affected. Thus, if the *TPMT* genotype is normal and the NUDT15 phenotype is an intermediate metabolizer, the dosing follows the recommendation for the NUDT15 intermediate metabolizer. When genotyping only *TPMT* (combined minor allele frequency of 5.6%) almost half of the individuals requiring dose adjustment based on the most up-to-date information are missed, while treating patients of Finnish ancestry.

The strength of our study is the large sample size, genotyped with a GWAS chip, which enabled us to estimate the LD structure in more detail, as compared with pharmacogenetic studies which have relied on genotyping of only a few variants. A limitation is that the sample was ascertained based on a previous diagnosis of psychosis and many patients were recruited from long-term treatment facilities and housing-units, which may skew the results towards patients who have had a sub-optimal therapeutic response, and thus divert the pharmacogenetic variant frequencies from the population mean. The BrePainGen study, from which the imputation panel was derived, included only women. However, this is not a problem because the *CYP2D6* gene is located in the autosome and the prediction of CYP2D6 phenotype from genotype does not differ between males and females^72^. The geographical difference in CYP2D6 and CYP2C19 phenotypes may also arise from trends in relocation instead of population history. Despite this, the geographical information can be used to inform local clinicians.

In conclusion, we demonstrate that despite complex structural variations in the *CYP2D6* locus, the copy-number can be computationally imputed. As the locus has consistently been shown to associate with drug concentrations, researchers are now able to expand these studies to disease outcomes using biobank data, for example. This will boost the development of individualized dosing algorithms and might eventually translate to improved patient care, especially in psychiatry. Over 20% of drugs listed by FDA as having pharmacogenetic information on their labels are used to treat psychiatric diseases. From these drugs, 69 % are metabolized through CYP2D6^73^. Additionally, we show that the bottle-neck effect contributes to the frequency of pharmacogenetic markers in Finland by demonstrating a high frequency of the decreased function variant *UGT1A1*6*. Further research should evaluate whether the observed phenomena affect the generalizability of the results of clinical trials across different ancestral groups.

## Supporting information

STROBE checklist

## Data Availability

The data is available from THL biobank when released from original study

## Acknowledgements

We thank BrePainGen and SUPER-Finland study participants.

---

**Supplemental Table 2.**
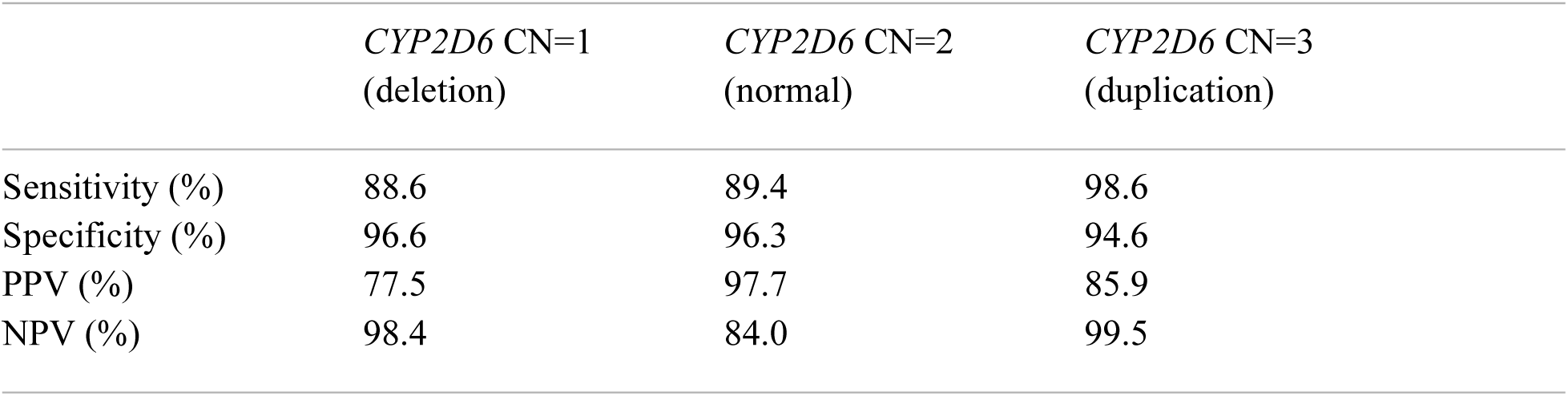
Sensitivity, specificity, positive predictive value (PPV) and negative predictive value (NPV) for *CYP2D6* copy-number (CN) in SUPER-Finland.

**Supplemental Table 3.** Distribution of CYP2D6 metabolizers by recruitment area in SUPER-Finland.

| % (n) | UM | NM | IM | PM | n total |
| --- | --- | --- | --- | --- | --- |
| Helsinki | 6.5 (152) | 61.2 (1426) | 28.4 (662) | 3.8 (89) | 2329 |
| Tampere | 7.5 (172) | 62.1 (1428) | 27.7 (637) | 2.7 (61) | 2298 |
| Kuopio | 5.0 (94) | 65.6 (1238) | 26.1 (493) | 3.2 (61) | 1886 |
| Oulu | 7.7 (134) | 61.5 (1067) | 27.8 (483) | 3.0 (52) | 1736 |
| Turku | 5.4 (55) | 64.3 (652) | 26.7 (270) | 3.6 (36) | 1013 |

UM, ultrarapid metabolizer; NM, normal metabolizer; IM, intermediate metabolizer; PM, poor metabolizer

**Supplemental table 4.** Distribution of CYP2C19 metabolizers by recruitment area in SUPER-Finland.

| % (n) | UM | RM | NM | IM | PM | n total |
| --- | --- | --- | --- | --- | --- | --- |
| Helsinki | 3.6 (83) | 24.0 (561) | 39.4 (917) | 29.0 (676) | 4.0 (92) | 2329 |
| Tampere | 4.1 (94) | 23.3 (535) | 41.1 (945) | 27.9 (641) | 3.6 (83) | 2298 |
| Kuopio | 4.0 (76) | 23.7 (447) | 40.2 (758) | 28.2 (532) | 3.9 (73) | 1886 |
| Oulu | 3.8 (65) | 25.6 (444) | 38.4 (668) | 29.6 (513) | 2.6 (46) | 1736 |
| Turku | 3.7 (37) | 26.3 (267) | 38.3 (388) | 29.0 (294) | 2.7 (27) | 1013 |

UM, ultrarapid metabolizer; RM, rapid metabolizer; NM, normal metabolizer; IM, intermediate metabolizer; PM, poor metabolizer.

## References

1. Wang, L., McLeod, H. L. & Weinshilboum, R. M. Genomics and drug response. New England Journal of Medicine 364, 1144–1153 (2011).

2. Pirmohamed, M. Personalized Pharmacogenomics: Predicting Efficacy and Adverse Drug Reactions. Annu. Rev. Genomics Hum. Genet. 15, 349–370 (2014).

3. Jukić, M. M., Haslemo, T., Molden, E. & Ingelman-Sundberg, M. Impact of CYP2C19 genotype on escitalopram exposure and therapeutic failure: A retrospective study based on 2,087 patients. Am. J. Psychiatry 175, 463–470 (2018).

4. Jukic, M. M., Smith, R. L., Haslemo, T., Molden, E. & Ingelman-Sundberg, M. Effect of CYP2D6 genotype on exposure and efficacy of risperidone and aripiprazole: a retrospective, cohort study. The Lancet Psychiatry 6, 418–426 (2019).

5. Tobias, J. D., Green, T. P. & Coté, C. J. Codeine: Time to say no. Pediatrics 138, 1–7 (2016).

6. Poulussen, F. C. P. et al. The effect of the CYP2D6 genotype on the maintenance dose of metoprolol in a chronic Dutch patient population. Pharmacogenet. Genomics 29, 179–182 (2019).

7. Li, S. et al. A meta-analysis of the effect of CYP2D6 polymorphism on the pharmacokinetics and pharmacodynamics of metoprolol. Int. J. Clin. Pharmacol. Ther. 55, 483–492 (2017).

8. Claassens, D. M. F. et al. A genotype-guided strategy for oral P2Y12 inhibitors in primary PCI. N. Engl. J. Med. 381, 1621–1631 (2019).

9. Zhou, S. F. Polymorphism of human cytochrome p450 2D6 and its clinical significance: Part I. Clinical Pharmacokinetics 48, 689–723 (2009).

10. Nofziger, C. et al. PharmVar GeneFocus: CYP2D6. Clinical Pharmacology and Therapeutics 107, 154–170 (2020).

11. Gaedigk, A., Dinh, J. C., Jeong, H., Prasad, B. & Leeder, J. S. Ten years’ experience with the CYP2D6 activity score: A perspective on future investigations to improve clinical predictions for precision therapeutics. Journal of Personalized Medicine 8, 1–15 (2018).

12. Aklillu, E. et al. Frequent distribution of ultrarapid metabolizers of debrisoquine in an ethiopian population carrying duplicated and multiduplicated functional CYP2D6 alleles. J. Pharmacol. Exp. Ther. 278, 441–446 (1996).

13. Bathum, L., Johansson, I., Ingelman-Sundberg, M., Hørder, M. & Brøsen, K. Ultrarapid metabolism of sparteine: Frequency of alleles with duplicated CYP2D6 genes in a Danish population as determined by restriction fragment length polymorphism and long polymerase chain reaction. Pharmacogenetics 8, 119–123 (1998).

14. Gaedigk, A., Sangkuhl, K., Whirl-Carrillo, M., Klein, T. & Steven Leeder, J. Prediction of CYP2D6 phenotype from genotype across world populations. Genet. Med. 19, 69–76 (2017).

15. Pietarinen, P., Tornio, A. & Niemi, M. High Frequency of CYP2D6 Ultrarapid Metabolizer Genotype in the Finnish Population. Basic Clin. Pharmacol. Toxicol. 119, 291–296 (2016).

16. Zhou, Y., Ingelman-Sundberg, M. & Lauschke, V. M. Worldwide Distribution of Cytochrome P450 Alleles: A Meta-analysis of Population-scale Sequencing Projects. Clin. Pharmacol. Ther. 102, 688–700 (2017).

17. Relling, Mary V., Evans, W. E. Pharmacogenomics in the clinic. Nature 526, 343–350 (2015).

18. Schuck, R. N. & Grillo, J. A. Pharmacogenomic biomarkers: An FDA perspective on utilization in biological product labeling. AAPS Journal 18, 573–577 (2016).

19. Ehmann, F. et al. Pharmacogenomic information in drug labels: European Medicines Agency perspective. Pharmacogenomics Journal 15, 201–210 (2015).

20. Keepers, G. A. et al. The American psychiatric association practice guideline for the treatment of patients with schizophrenia. Am. J. Psychiatry 177, 868–872 (2020).

21. Johnson, J. A. Ethnic differences in cardiovascular drug response: Potential contribution of pharmacogenetics. Circulation 118, 1383–1393 (2008).

22. Marchini, J. & Howie, B. Genotype imputation for genome-wide association studies. Nat. Rev. Genet. 11, 499–511 (2010).

23. Tewhey, R., Bansal, V., Torkamani, A., Topol, E. J. & Schork, N. J. The importance of phase information for human genomics. Nat. Genet. 12, 215–223 (2011).

24. Caspar, S. M., Schneider, T., Meienberg, J. & Matyas, G. Added Value of Clinical Sequencing: WGS-Based Profiling of Pharmacogenes. Int. J. Mol. Sci. 21, 2308 (2020).

25. Lim, E. T. et al. Distribution and medical impact of loss-of-function variants in the Finnish founder population. PLoS Genet. 10, 1–12 (2014).

26. Chheda, H. et al. Whole-genome view of the consequences of a population bottleneck using 2926 genome sequences from Finland and United Kingdom. Eur. J. Hum. Genet. 25, 477–484 (2017).

27. Norio, R. Finnish Disease Heritage I: Characteristics, causes, background. Human Genetics 112, 441–456 (2003).

28. Norio, R. Finnish Disease Heritage II: Population prehistory and genetic roots of Finns. Human Genetics 112, 457–469 (2003).

29. Sund, R. Quality of the Finnish Hospital Discharge Register: A systematic review. Scand. J. Public Health 40, 505–515 (2012).

30. Mitt, M. et al. Improved imputation accuracy of rare and low-frequency variants using population-specific high-coverage WGS-based imputation reference panel. Eur. J. Hum. Genet. 25, 869–876 (2017).

31. Kaunisto, M. A. et al. Pain in 1,000 women treated for breast cancer: a prospective study of pain sensitivity and postoperative pain. Anesthesiology 119, 1410–1421 (2013).

32. Cajanus, K. et al. Analgesic Plasma Concentrations of Oxycodone After Surgery for Breast Cancer—Which Factors Matter? Clin. Pharmacol. Ther. 103, 653–662 (2018).

33. Loh, P.-R. et al. Reference-based phasing using the Haplotype Reference Consortium panel. Nat. Genet. 48, 1443–1448 (2016).

34. Loh, P.-R., Palamara, P. F. & Price, A. L. Fast and accurate long-range phasing in a UK Biobank cohort. Nat. Publ. Gr. 48, 811–819 (2016).

35. Browning, B. L. & Browning, S. R. Genotype Imputation with Millions of Reference Samples. Am. J. Hum. Genet. 98, 116–126 (2016).

36. Karnes, J. H. et al. Clinical Pharmacogenetics Implementation Consortium (CPIC) Guideline for CYP2C9 and HLA-B Genotypes and Phenytoin Dosing: 2020 Update. Clin. Pharmacol. Ther. (2020). doi:10.1002/cpt.2008

37. Hicks, J. K. et al. Clinical Pharmacogenetics Implementation Consortium (CPIC) guideline for CYP2D6 and CYP2C19 genotypes and dosing of selective serotonin reuptake inhibitors. Clin. Pharmacol. Ther. 98, 127–134 (2015).

38. Crews, K. R. et al. Clinical pharmacogenetics implementation consortium guidelines for cytochrome P450 2D6 genotype and codeine therapy: 2014 Update. Clin. Pharmacol. Ther. 95, 376–382 (2014).

39. Amstutz, U. et al. Clinical Pharmacogenetics Implementation Consortium (CPIC) Guideline for Dihydropyrimidine Dehydrogenase Genotype and Fluoropyrimidine Dosing: 2017 Update. Clin. Pharmacol. Ther. 103, 210–216 (2018).

40. Relling, M. V et al. Clinical Pharmacogenetics Implementation Consortium Guideline for Thiopurine Dosing Based on TPMT and NUDT15 Genotypes: 2018 Update. Clin. Pharmacol. Ther. 105, 1095–1105 (2019).

41. Ramsey, L. et al. The Clinical Pharmacogenetics Implementation Consortium Guideline for SLCO1B1 and Simvastatin-Induced Myopathy: 2014 Update. Clin. Pharmacol. Ther. 1–6 (2014). doi:10.1038/clpt.2014.125

42. Gammal, R. S. et al. Clinical Pharmacogenetics Implementation Consortium (CPIC) Guideline for UGT1A1 and Atazanavir Prescribing. Clin. Pharmacol. Ther. 00, 1–7 (2015).

43. Carter, J. V, Pan, J., Rai, S. N. & Galandiuk, S. Education ROC-ing along: Evaluation and interpretation of receiver operating characteristic curves. Surgery 159, 1638–1645 (2016).

44. R Core Team. R: A language and environment for statistical computing. R Foundation for Statistical Computing, Vienna, Austria. (2018).

45. Karczewski, K. J. et al. The mutational constraint spectrum quantified from variation in 141,456 humans. Nature 581, 434–443 (2020).

46. Kelley, M. E. et al. Response rate profiles for major depressive disorder: Characterizing early response and longitudinal nonresponse. Depress. Anxiety 35, 992–1000 (2018).

47. Heise, T. et al. Pharmacodynamic Effects of Single and Multiple Doses of Empagliflozin in Patients With Type 2 Diabetes. Clin. Ther. 38, 2265–2276 (2016).

48. Gelenberg, A. J. et al. Practice Guideline for the Treatment of Patients With Major Depressive Disorder. American Psychiatric Association (2010).

49. Bradley, P. et al. Improved efficacy with targeted pharmacogenetic-guided treatment of patients with depression and anxiety: A randomized clinical trial demonstrating clinical utility. J. Psychiatr. Res. 96, 100–107 (2018).

50. Ripke, S. et al. Biological insights from 108 schizophrenia-associated genetic loci. Nature 511, 421–427 (2014).

51. Smith, R. L. et al. Identification of a novel polymorphism associated with reduced clozapine concentration in schizophrenia patients—a genome-wide association study adjusting for smoking habits. Transl. Psychiatry 10, 1–10 (2020).

52. Meade, T. et al. SLCO1B1 variants and statin-induced myopathy - A genomewide study. N. Engl. J. Med. 359, 789–799 (2008).

53. Yang, S. K. et al. A common missense variant in NUDT15 confers susceptibility to thiopurine-induced leukopenia. Nat. Genet. 46, 1017–1020 (2014).

54. Krebs, K. & Milani, L. Translating pharmacogenomics into clinical decisions: do not let the perfect be the enemy of the good. Human genomics 13, 1–13 (2019).

55. Huang, L., Wang, C. & Rosenberg, N. A. The Relationship between Imputation Error and Statistical Power in Genetic Association Studies in Diverse Populations. Am. J. Hum. Genet. 85, 692–698 (2009).

56. Weedon, M. et al. Very rare pathogenic genetic variants detected by SNP-chips are usually false positives: implications for direct-to-consumer genetic testing. bioRxiv (2019). doi:10.1101/696799

57. Jackson, L. et al. Assessing the analytical validity of SNP-chips for detecting very rare pathogenic variants: implications for direct-to-consumer genetic testing. bioRxiv (2019). doi:10.1101/696799

58. Sistonen, J. et al. Pharmacogenetic variation at CYP2C9, CYP2C19, and CYP2D6 at global and microgeographic scales. Pharmacogenet. Genomics 19, 170–179 (2009).

59. Hikino, K. et al. Comparison of effects of UGT1A1*6 and UGT1A1*28 on irinotecan-induced adverse reactions in the Japanese population: analysis of the Biobank Japan Project. J. Hum. Genet. 64, 1195–1202 (2019).

60. Hirvensalo, P. et al. UGT1A3 and Sex Are Major Determinants of Telmisartan Pharmacokinetics—A Comprehensive Pharmacogenomic Study. Clin. Pharmacol. Ther. 0, 1–11 (2020).

61. Takano, M. & Sugiyama, T. UGT1A1 polymorphisms in cancer: Impact on irinotecan treatment. Pharmacogenomics and Personalized Medicine 10, 61–68 (2017).

62. Kadakol, A. et al. Genetic lesions of bilirubin uridine-diphosphoglucuronate glucuronosyltransferase (UGT1A1) causing Crigler-Najjar and Gilbert syndromes: Correlation of genotype to phenotype. Human Mutation 16, 297–306 (2000).

63. Bosma, P. J. et al. The genetic basis of the reduced expression of bilirubin udp-glucuronosyltransferase 1 in gilbert’s syndrome. N. Engl. J. Med. 333, 1171–1175 (1995).

64. Machiela, M. J. & Chanock, S. J. LDlink: a web-based application for exploring population-specific haplotype structure and linking correlated alleles of possible functional variants. Bioinformatics 31, 3555–3557 (2015).

65. Hicks, J. K. et al. Clinical pharmacogenetics implementation consortium guideline (CPIC) for CYP2D6 and CYP2C19 genotypes and dosing of tricyclic antidepressants: 2016 update. Clin. Pharmacol. Ther. 102, 37–44 (2017).

66. Scott, S. A. et al. Clinical pharmacogenetics implementation consortium guidelines for CYP2C19 genotype and clopidogrel therapy: 2013 update. Clin. Pharmacol. Ther. 94, 317–323 (2013).

67. Lima, J. J. et al. Clinical Pharmacogenetics Implementation Consortium (CPIC) Guideline for CYP2C19 and Proton Pump Inhibitor Dosing. Clin. Pharmacol. Ther. 0, 1–7 (2020).

68. Mega, J. L. et al. Reduced-function CYP2C19 genotype and risk of adverse clinical outcomes among patients treated with clopidogrel predominantly for PCI: A meta-analysis. JAMA -J. Am. Med. Assoc. 304, 1821–1830 (2010).

69. Brown, S. A. & Pereira, N. Pharmacogenomic impact of CYP2C19 variation on clopidogrel therapy in precision cardiovascular medicine. Journal of Personalized Medicine 8, 1–31 (2018).

70. Kelley, K. J. Siponimod Approved to Treat Relapsing Forms of MS. NEJM J. Watch (2019). doi:10.1056/NEJM-JW.FW115220

71. Johnson, J. A. et al. Clinical Pharmacogenetics Implementation Consortium (CPIC) Guideline for Pharmacogenetics-Guided Warfarin Dosing: 2017 Update. Clin. Pharmacol. Ther. 102, 397–404 (2017).

72. Gaedigk, A. et al. The CYP2D6 Activity Score: Translating Genotype Information into a Qualitative Measure of Phenotype. Clin. Pharmacol. Ther. 83, (2008).

73. Jarvis, J. P., Peter, A. P. & Shaman, J. A. Consequences of CYP2D6 Copy-Number Variation for Pharmacogenomics in Psychiatry. Frontiers in Psychiatry 10, (2019).

